# Identification of a presymptomatic and early disease signature for Amyotrophic Lateral Sclerosis (ALS): protocol of the premodiALS study

**DOI:** 10.1101/2025.05.27.25328387

**Authors:** Laura Tzeplaeff, Ana Galhoz, Clara Meijs, Lucas Caldi Gomes, Andrej Kovac, Amrei Menzel, Hatice Değirmenci, Abir Alaamel, Hüseyin Can Kaya, Ali Günalp Çelik, Sine Dinçer, Meltem Korucuk, Sibel Berker Karaüzüm, Elif Bayraktar, Vildan Çiftçi, Uğur Bilge, Filiz Koç, Antonia F Demleitner, Anne Buchberger, Ricarda von Heynitz, Vincent Gmeiner, Christina Knellwolf, Mohammed Mouzouri, Joanne Wuu, A. Nazlı Başak, Peter Munch Andersen, Florian Kohlmayer, Nicholas J Ashton, Wojciech Kuban, Christof Lenz, Mary-Louise Rogers, Norbert Zilka, Philippe Corcia, Yossef Lerner, Markus Weber, Monika Turcanova Koprusakova, Hilmi Uysal, Michael Benatar, Michael P Menden, Paul Lingor

**Author notes:** Corresponding Author: Laura Tzeplaeff, Department of Neurology, Klinikum rechts der Isar, TUM Universitätsklinikum, School of Medicine and Health, Technical University of Munich, Ismaninger Str. 22, 81675 Munich.

## Abstract

The median time to diagnosis of amyotrophic lateral sclerosis (ALS) is approximately 12 months after the onset of first symptoms. This diagnostic delay is primarily due to the nonspecific nature of early symptoms and the clinical challenges in differentiating ALS from its mimics. Therefore, the discovery of reliable biomarkers for the early and accurate diagnosis of ALS represents a critical medical need. A total of 330 participants will be recruited across six international study sites. The cohort will include (1) pre-symptomatic gene mutation carriers, (2) symptomatic individuals up to 12 months after symptom onset with either ALS, ALS mimics, or a pure motor syndrome with yet unclear assignment, and (3) healthy controls. Participants will engage in a one-year longitudinal study, consisting of an initial evaluation at baseline visit and a follow-up visit 12 months later. Assessments will include an environmental and medical history questionnaire, neurological examinations, olfactory testing, cognitive/behavioral evaluations, and the collection of biological samples (serum, plasma, urine, tear fluid, and cerebrospinal fluid). Proteomic, metabolomic, and lipidomic analyses will be performed using mass spectrometry and targeted immunoassays, with all samples processed under standardized protocols. The resulting multimodal dataset will be systematically integrated in an effort to uncover a clinico-molecular signature characteristic of presymptomatic and early ALS. These findings may have relevance to early ALS diagnosis and future clinical practice.

## Introduction

Amyotrophic lateral sclerosis (ALS) is the most common adult-onset motor neuron disease. Despite advanced healthcare systems in industrialized countries, the average diagnostic delay for ALS remains approximately 12 months after symptom onset [1–3]. This delay is particularly critical given the median life expectancy of only 3 to 5 years post-symptom onset. Moreover, nearly half of ALS patients are initially misdiagnosed, largely due to limited familiarity with ALS among general practitioners and to the clinical challenges in distinguishing ALS from other disorders that mimic its early symptoms [1, 2, 4]. Given the rapid progression, the identification of reliable biomarkers for early and accurate diagnosis is essential to initiate timely therapy and facilitate patient enrollment in clinical trials.

Biomarker research in ALS has identified some promising candidates with potential for improving diagnosis and disease monitoring. Neurofilaments (Nf) are the most extensively studied biomarkers for ALS. Both phosphorylated Nf heavy chain (pNfH) and Nf light chain (NfL) levels in serum and cerebrospinal fluid (CSF) are increased in ALS compared to healthy controls and mimics, correlating with the rate of neuronal and axonal damage [5, 6]. Recently, NfL was also shown to be a useful marker of therapy response [7]. Additionally, urinary molecules have been studied as markers of disease progression, including the soluble extracellular domain of the neurotrophin receptor p75 (sp75^ECD^) [8, 9] and neopterin [10]. Other proteins that have been identified to be differentially regulated in ALS cohorts include chitinases (CHIT), chitinase-like proteins (CHITL), creatine kinase (CK), troponin (TnT), total Tau and p-Tau, collagen proteins, and several inflammatory markers such as MCP-1, IL-6, and IL-18 [11, 12]. In addition to blood- or CSF-derived molecules, tear fluid may hold potential for biomarker discovery, as demonstrated in other neurodegenerative disorders [13, 14].

Biomarkers have the potential to predict the timing of phenoconversion or identify sporadic patients in the presymptomatic phase of ALS, which refers to the period before the clinical manifestation of the disease [15–17]. Although the majority of ALS cases are sporadic (sALS), approximately 10-15% can be attributed to a genetic cause, even in the absence of a family history (genetic ALS or gALS) [18, 19]. However, almost half of gALS cases show a positive family history (fALS). In 50-70% of fALS cases, the cause of disease can be attributed to known mutations, such as repeat expansions in the *C9orf72* gene or mutations in the *SOD1*, *TARDBP* and *FUS* genes [19]. Genetic testing of biological relatives of fALS cases can identify individuals who carry ALS-associated pathogenic variants but have not yet developed clinical manifestations of the disease, hereafter referred to as "pre-symptomatic gene mutation carriers" (PGMC).

Our hypothesis is that PGMC gradually accumulate molecular changes that eventually lead to the clinical manifestation of ALS. Previous studies demonstrated significant alterations in PGMC well before the manifestation of symptoms. For instance, NfL and pNfH are elevated in the serum and CSF of PGMC prior to phenoconversion [20, 21]. Notably, elevated NfL levels have also been observed in non-mutation carriers who were later diagnosed with ALS, suggesting that NfL could serve as a susceptibility/risk marker in the early stages of the disease [22, 23]. More recent studies have suggested that cryptic Hepatoma-derived growth Factor-Like Protein 2 (HDGFL2) peptides resulting from cryptic splicing may be detected in *C9orf72* PGMC even earlier than NfL, but limited longitudinal data and an absence of data from phenoconverters indicates the need for further studies [24, 25].

Alterations in cognition (e.g. verbal fluency), neuroimaging (e.g. volumetric changes in specific anatomical regions), electrophysiology, sleep (e.g. polysomnography), metabolic (e.g. resting metabolic rate) and proteomics have been observed in PGMC before symptom onset, highlighting early ALS pathophysiology [26–34]. Although group level differences may inform about disease mechanisms, they may not be suitable as susceptibility/risk or diagnostic markers. Therefore, biomarkers that operate at the level of an individual and predict susceptibility to developing clinically manifest disease are needed. The identification of disease biomarkers that precede axonal loss could help to explore their utility as diagnostic marker in patients in whom the ALS diagnosis is unclear. Such discovery could significantly transform ALS diagnosis and treatment. Pre-symptomatic clinical assessments and multi-omic biofluid profiling offer promising avenues for uncovering novel biomarker combinations, as seen in other neurodegenerative diseases like Alzheimer’s and Parkinson’s disease [35–38]. These approaches could enable the identification of individuals at risk when no genetic cause is identified, predict phenoconversion, provide an earlier diagnosis and finally improve access to clinical trials and treatments in ALS.

Here, we present the design of the premodiALS study, an international project focused on identifying a clinico-molecular signature that differentiates PGMC from healthy controls and ALS from ALS mimics, ultimately contributing to a deeper understanding of ALS pathophysiology.

## Material and Methods

### Objectives and study design

The premodiALS study is a multicenter, prospective investigation aimed at identifying clinical markers and molecular biomarkers, and their combinations, that differentiate PGMC and early ALS cases from healthy controls and ALS mimics. Participants are assessed longitudinally at two time points: a baseline visit (V0) and a follow-up visit (V1) at an interval of one year (**Fig. 1**).

**Fig. 1.**
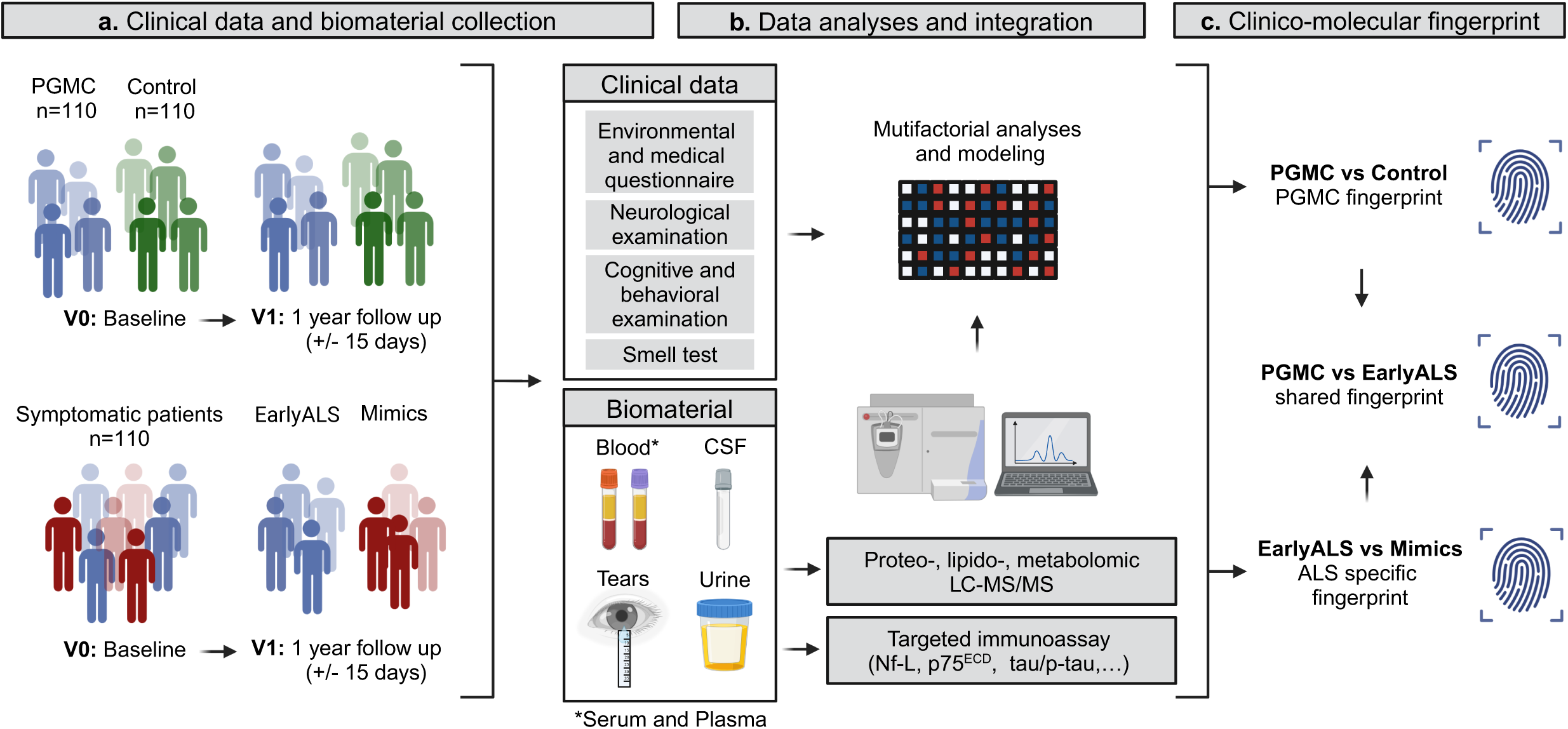
Overview of the premodiALS study procedures, analyses and outcomes. (a) Clinical and biomaterial collection from the PGMC, control and symptomatic patients’ group. The symptomatic patients group includes subjects within 12 months of symptom onset with either early ALS or an ALS mimic, or a pure motor syndrome that does not permit to establish a certain diagnosis at the baseline visit (V0). The categorization into early ALS and ALS mimic groups will be determined *a posteriori* at the follow up visit (V1). (b) Analysis and integration of the clinical data and molecular data obtained from the biomaterial collection. (c) Identification of a clinico-molecular fingerprint of PGMC and early ALS. V: visit; PGMC: pre-symptomatic gene mutation carrier. *Created in BioRender. Tzeplaeff, L. (2025)* https://BioRender.com/v87e030

### Participants

#### Recruitment

Recruitment started in the 1st quarter of 2023 and is still ongoing. Patients are recruited at 6 international study sites: Klinikum rechts der Isar of the Technical University of Munich, Munich, Germany; Akdeniz University Hospital, Antalya, Turkiye; Hadassah Ein Karem University Hospital, Jerusalem, Israel; Kantonsspital St.Gallen, St. Gallen, Switzerland; University Hospital Martin, Slovakia; and The CHRU of Tours, Tours, France. All recruitment centers are well-known ALS reference centers. The recruitment goal is a total of 110 PGMC, 110 control and 110 pure motor syndrome/early ALS/ALS mimic patients.

### Participant group description

Three groups of participants are included: (1) Pre-symptomatic gene mutation carriers (PGMC), (2) symptomatic subjects including either patients with a pure motor syndrome or early ALS or ALS mimics, and (3) controls (CTR).

1. PGMC are individuals who carry a pathogenic variant in a known ALS-associated gene but have not yet developed clinically manifest ALS. Genetic testing must have been conducted by a clinically certified laboratory.
2. The second group consists of symptomatic patients (SYMP) within 12 months after symptom onset. These may present either as early ALS (as per the El Escorial criteria [39]) or as known ALS mimic or as patients with a pure motor syndrome, which is insufficient to establish an ALS diagnosis at the baseline visit (V0), and who may later develop into either ALS or an ALS mimic. ALS mimics refer to diseases that initially exhibit clinical symptoms resembling ALS, such as multifocal motor neuropathy, benign fasciculations, myopathies/myositides, radiculopathies, generalized myasthenia gravis, and others. The definite classification into “early ALS” or “ALS mimic” will be established at the follow-up visit (V1), when clinical progression provides diagnostic certainty.
3. Control (CTR) subjects can be family members of PGMC who are not carrying the genetic mutation, as well as life partners, spouses or friends, who are exposed to similar environmental factors, are within a comparable age range and do not show any motor symptoms.

### Inclusion and exclusion criteria

All groups include male and female subjects aged 18 years or older, provided that informed consent to participate in the study is obtained. Exclusion criteria include individuals who do not consent to biosampling or are unable to attend the V1 follow-up visit (12 months after baseline) at the study center. Subjects on anticoagulation therapy may participate, but they will be excluded from lumbar puncture.

### Study procedures

#### Visit plan and assessments

Informed consent is obtained before any study-specific procedures are performed. A total of two visits (V0 and V1) at an interval of one year (+/- 15 days) are performed (**Table 1**). During the baseline V0, a questionnaire focused on demographics, environmental and medical history, as well as the ALS Functional Rating Scale - Revised (ALSFRS-R), is completed by the participant together with a trained study nurse or research coordinator to ensure that all questions are answered to avoid missing data (**Supplemental Files 1)**. For ALSFRS-R assessment, translated versions of the scale were used in each country, some of which have been validated previously [40–42]. The baseline V0 assessment includes a detailed neurological examination; a cognitive and behavioral examination using the Edinburgh Cognitive and Behavioral ALS Screen (ECAS), including the caregiver/informant interview to detect behavioral abnormalities common in behavioral variant of FTD; as well as a standardized brief smell identification test (B-SIT). The questionnaire (**Supplemental Files 1)** and ECAS are provided in the participant’s native language (i.e., German, Swiss-German, French, Slovak, Turkish, and Hebrew). Officially translated and culturally adapted version of the ECAS exist for all of these languages, except Turkish, which is currently adapted in the scope of the premodiALS study. The B-SIT is provided in either German, French, Turkish or English. To account for potential olfactory disturbances caused by SARS-CoV-2 infection, participants are asked about their history of SARS-CoV-2 infection and whether they have experienced any issues with their sense of smell. The collected biological samples include blood (serum, plasma), cerebrospinal fluid (CSF), urine and tear fluid (collected with Schirmer test strips). Before CSF collection, coagulation parameters (e.g. the international normalized ratio (INR), the partial thromboplastin time (PTT) and platelet count) are analyzed. Participants with an altered coagulation state will be excluded from the lumbar puncture procedure, but will not be excluded from the study.

**Table 1.**
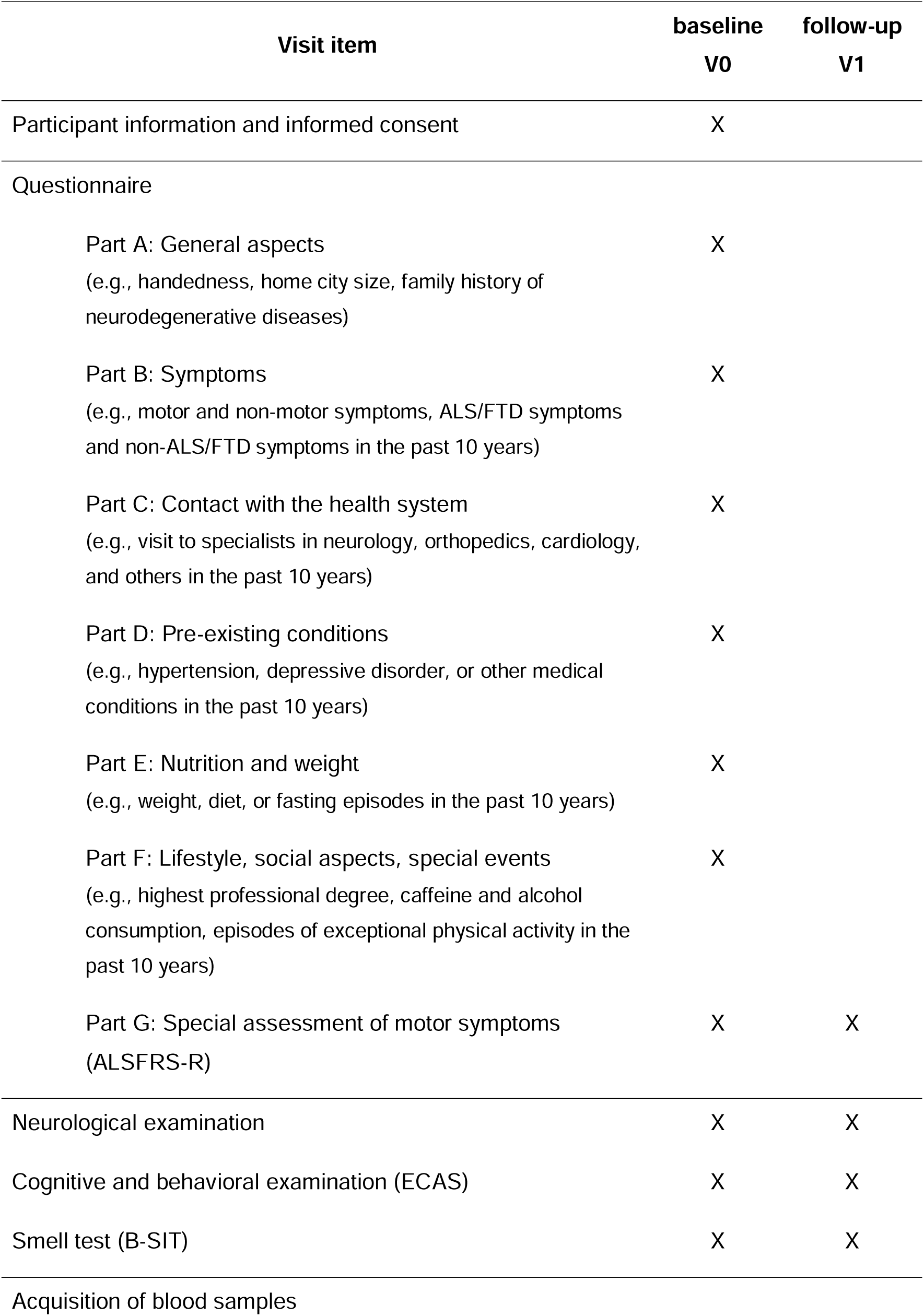
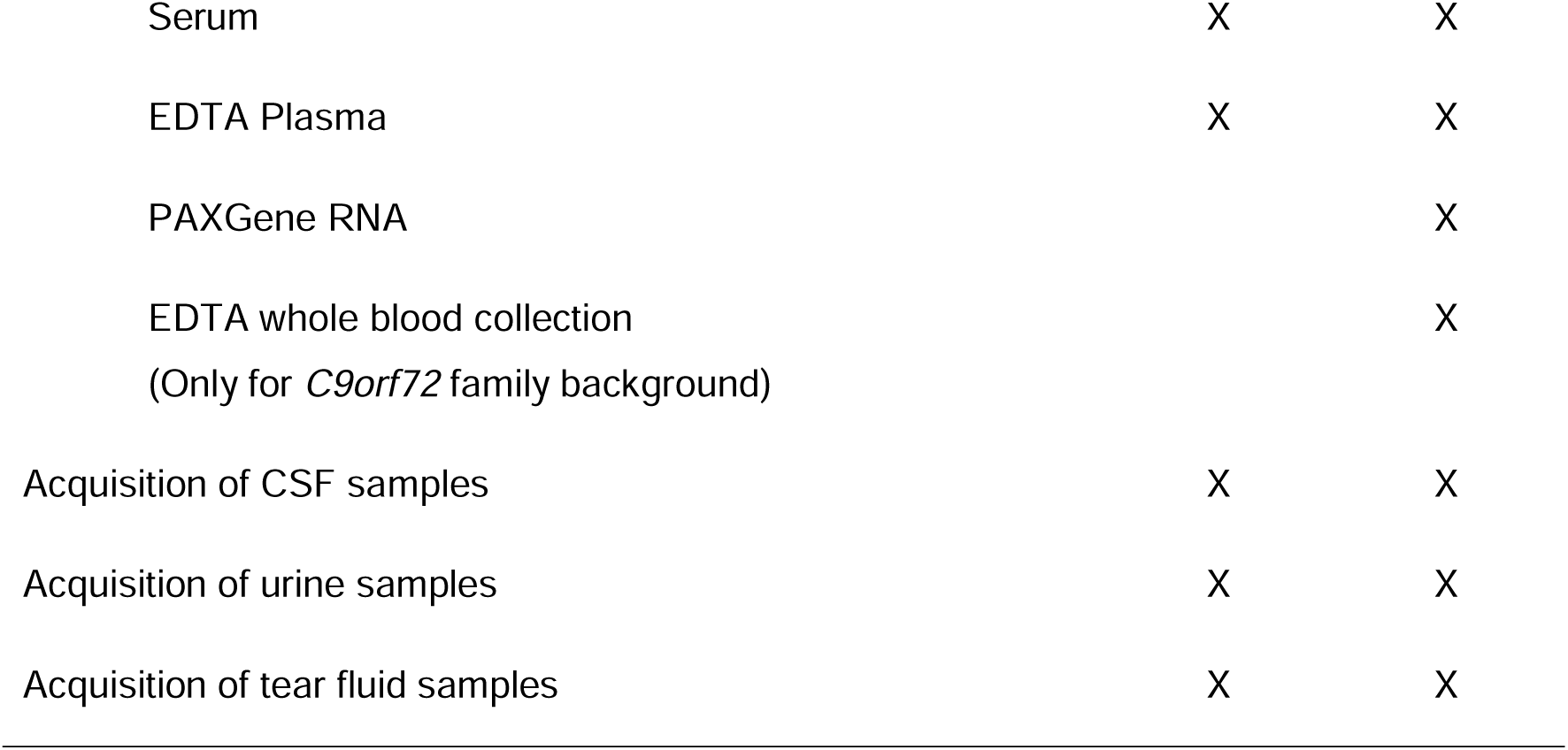
Study visit schedule.

| Visit item | baseline<br>V0 | follow-up<br>V1 |
| --- | --- | --- |
| Participant information and informed consent | X |  |
| Questionnaire |  |  |
| Part A: General aspects<br>(e.g., handedness, home city size, family history of neurodegenerative diseases) | X |  |
| Part B: Symptoms<br>(e.g., motor and non-motor symptoms, ALS/FTD symptoms and non-ALS/FTD symptoms in the past 10 years) | X |  |
| Part C: Contact with the health system<br>(e.g., visit to specialists in neurology, orthopedics, cardiology, and others in the past 10 years) | X |  |
| Part D: Pre-existing conditions<br>(e.g., hypertension, depressive disorder, or other medical conditions in the past 10 years) | X |  |
| Part E: Nutrition and weight<br>(e.g., weight, diet, or fasting episodes in the past 10 years) | X |  |
| Part F: Lifestyle, social aspects, special events<br>(e.g., highest professional degree, caffeine and alcohol consumption, episodes of exceptional physical activity in the past 10 years) | X |  |
| Part G: Special assessment of motor symptoms<br>(ALSFRS-R) | X | X |
| Neurological examination | X | X |
| Cognitive and behavioral examination (ECAS) | X | X |
| Smell test (B-SIT) | X | X |
| Acquisition of blood samples |  |  |
| Serum | X | X |
| EDTA Plasma | X | X |
| PAXGene RNA |  | X |
| EDTA whole blood collection<br>(Only for *C9orf72* family background) |  | X |
| Acquisition of CSF samples | X | X |
| Acquisition of urine samples | X | X |
| Acquisition of tear fluid samples | X | X |

During the follow-up V1, all assessments are repeated in the same way as for the baseline V0, with the exception of the questionnaire, where only the ALSFRS-R is repeatedly assessed.

In addition to the predefined sample collection, each participant may consent to donate additional biomaterial for biobanking during both baseline V0 and follow-up V1. This additional material is obtained simultaneously with the predefined biomaterial study collection and will be available for future collaborative projects beyond what is specified in the premodiALS protocol.

### Biomaterial collection protocol

All biomaterial samples are collected on ice, then processed and frozen at -80°C as quickly as possible, within 30 to 60 minutes after collection.

Blood collection: During each visit, venous blood is drawn in 2 x 10 mL clot activator tube (CAT) serum vacutainers (BD, 367896) for serum collection and in 2 x 10 mL EDTA K2 vacutainers (BD, 367525) for plasma collection. After collection, tubes are gently inverted 10 times, centrifuged at ≤1300 g for 10 minutes at 18–25°C. Supernatants are then aliquoted into 12 tubes of 500 µL each, and stored at -80°C until further use of the biomaterial. In addition, 3 mL of blood is collected into a citrate tube and 2.6 mL of blood is collected in a EDTA monovette tube. These two tubes of blood are shipped to the local laboratory to determine coagulation parameters (INR, PTT, and platelets) before lumbar puncture for CSF collection. During the follow up visit (V1), 1 x 2.5 mL of blood is collected in a PAXgene RNA tube (BD, 762165) for isolation and purification of intracellular RNA. Tubes are inverted 10 times, kept in an upright position at room temperature for 2 hours, transferred first to a -20°C freezer for approximately 24 hours as required by the manufacturer, and finally transferred to a -80°C freezer, and kept at this temperature until shipment. If participants belong to a family with a *C9orf72* expansion genetic background, two tubes of 7.5 mL EDTA for whole blood will be additionally collected during the follow up visit (V1) and stored at -80°C.

CSF collection: After verification that the participant is not taking any anticoagulants and an unremarkable coagulation test the lumbar puncture is performed and 15 mL of CSF are collected in 2 polypropylene tubes. One tube is centrifuged at 2000 g for 10 minutes at 4°C, then supernatant is aliquoted into 14 tubes of 500 μL and kept at -80°C until further usage. The second tube is sent to the local laboratory for basic CSF examination to quantify white cell counts, lactate, total protein and to exclude blood contamination.

Urine collection: During each visit, participants are asked to dispense approximately 50 mL of urine into a urinalysis container. A urinalysis strip (UriScan™) is used to assess blood, bilirubin, urobilinogen, ketone, protein, nitrite, glucose, specific gravity, leukocytes and pH. From the urinalysis container, 15 mL of urine is collected in a 15 mL tube and centrifuged at 2000 g for 10 min at 4°C. Supernatant is then distributed into 8 aliquots of 500 μL and stored at -80°C until further usage.

Tear fluid collection: Schirmer test strips (MDT, TearFlo™) are inserted at the medial and lateral thirds of the lower eyelid of each eye. Collection lasts 5 minutes with eyes closed [43]. As the tear volume highly varies between individuals, the wetting length of each stripe is documented. Indeed, different wetting lengths can result in different protein concentrations and therefore impact analyses if not normalized. Stripes are individually placed in 500 μL tubes and stored at -80°C until usage.

### Outcome measures

#### Clinical data collection

Clinical data will be obtained through the neurological examination, the ECAS, the B- SIT, and the questionnaire. The neurological examination will include the following parameters: 1) cranial nerve status, tongue motility, dysarthria, dysphagia, 2) examination of the motor system with degrees of strength for neck flexion and extension, upper and lower extremities as well as muscle bulk and muscle tone, 3) testing of the deep tendon reflexes, the palmomental reflex, pathological reflexes, 4) examination of the sensory system, and 5) examination of the coordination, stance and gait. The B-SIT is composed of odors specific to the culture of the associated country language. The type and order of odors can therefore vary for different recruitment centers. For this reason, B-SIT data will be collected as a score of successes and errors on a total of 12 different items. The premodiALS questionnaire is divided in 7 Parts (Parts A-G) and includes the following parameters: A) general aspects, B) symptoms, C) contact with the health system, D) pre-existing conditions, E) nutrition and weight, F) lifestyle, social aspects, special events, and G) assessment of motor symptoms (ALSFRS-R). The full questionnaire is available in **Supplemental Files 1**.

### Molecular analyses

The biomaterial from both visits will be subjected to proteomic, metabolomic and lipidomic profiling, as well as the examination of individual disease-relevant proteins (**Table 2**). These data will be generated in five analytical centers. Plasma/serum and CSF proteome analyses by high resolution mass spectrometry will be performed at the Maj Institute of Pharmacology, Warsaw, Poland. The tear fluid proteome will be analyzed by high resolution mass spectrometry at the University Medical Center Göttingen, Göttingen, Germany. Metabolomic and lipidomic data will be obtained from plasma, urine and CSF via liquid chromatography tandem mass spectrometry (LC-MS/MS) at the Institute of Neuroimmunology, Slovak Academy of Sciences, Bratislava, Slovakia. Targeted immunoassays will be used for the examination of individual disease-relevant proteins. In urine samples, soluble neurotrophin receptor p75 extracellular domain (p75ECD) and neopterin will be analyzed with an enzyme-linked immunosorbent assay (ELISA) at the Flinders University, Adelaide, South Australia. Plasma/serum and CSF samples will be subjected to measurements of Nf-L, GFAP, total-Tau/phospho-Tau, as well as a panel of synaptic, lysosomal and inflammatory proteins using the digital ELISA based on single molecule arrays (SIMOA) and the Nucleic acid Linked Immuno-Sandwich Assay (NULISA) at the Sahlgrenska Academy at Gothenburg University, Mölndal Sweden.

**Table 2.** Summary of expected molecular outcome measures.

| Source sample type | Proteomics | Metabolomics/<br>lipidomics | Targeted<br>biomarkers |
| --- | --- | --- | --- |
| Blood |  |  |  |
| Plasma | X | X | X |
| Serum | X |  | X |
| Urine |  | X | X |
| CSF | X | X | X |
| Tear fluid | X |  |  |

Additional biomaterial (serum, plasma, CSF and urine) stored in the biobank and not yet allocated to predefined analyses will be available for collaborative projects.

### Statistical analyses

To identify a clinico-molecular fingerprint, we will initially analyze each dataset separately. In a subsequent step, we will integrate the relevant features to increase statistical power to identify robust signatures. Longitudinal difference between the baseline V0 and the follow-up V1 visit will be assessed for each group. Furthermore, we will systematically compare PGMC vs. CTR and early ALS vs. ALS mimics, adjusting for sex and age. The fingerprint of PGMC will also be compared with early ALS data to determine changes common to both groups. To describe the potentially strongest contrast, we will also compare early ALS and CTR groups. Where applicable, data will be stratified by sex and/or genetic mutation, or adjusted for these variables.

Clinical data will be comprehensively analyzed using both univariate and multivariate statistical methods to identify disease-specific clinical features. In the univariate framework, continuous and categorical variables from the questionnaire will be tested separately. Analyses will first be performed on the total data set with sex and age adjustment. In parallel, data will be stratified by sex and results will be reported using sex-adjusted odds ratio. Significance will be evaluated using Fisher’s exact test for categorical variables and the Wald test for continuous variables, respectively. Additionally, multivariate generalized linear models (GLM) will be employed to predict PGMC and early ALS participants, with sequential extraction of predictive features. Statistical findings will be corrected for multiple hypothesis testing utilizing the Benjamini-Hochberg (BH) correction at 5% False Discovery Rate (FDR).

Molecular data will be analyzed to identify differentially enriched proteins and metabolites. In concordance with clinical data analysis, the same 5% FDR cut-off will be employed. Gene Set Enrichment Analysis (GSEA) will be used for enrichment analysis with Gene Ontology (GO) annotations via the clusterProfiler package (v4.6.2) [44–46]. Given the heterogeneous nature of ALS, we will apply data-driven clustering methods and assess robustness with Akaike’s Information Criterion, Silhouette coefficient, and Dunn index.

Results from the metabolomic and proteomic analysis will be compared with results from the MAXOMOD consortium [47, 48]. Additionally, we will use the Wppi Bioconductor package to identify novel ALS-relevant proteins by constructing a protein-protein interaction (PPI) using Omnipath, which will be explored via a random walk with restart algorithm to identify new biomarker candidates [49].

We will apply Gaussian mixture models, spectral bi-clustering and Multi□Omics Factor Analysis (MOFA) to integrate clinical and molecular data to derive a robust presymptomatic and early ALS clinical-molecular fingerprint [50]. Key features of this fingerprint will be replicated in an independent PGMC cohort from the *Pre-fALS* study (University of Miami, Miami, FL, USA) [20]. In addition, supervised machine learning models (i.e., linear regressions, random forest, and deep learning models) will be employed to predict disease progression, mitigating the risk of overlooking critical biomarkers. These models will be cross-validated, and predictive features will be systematically identified and assessed.

## Results

### Ethics

This observational clinical study is conducted in accordance with the current ICH-GCP-guidelines and the Declaration of Helsinki. The studies involving humans are approved by the ethics committee at TU München [2022-520_2-S-SB] and are conducted following the local legislation and institutional requirements. We obtained ethics committee approval from all recruitment centers. All participants provided their written informed consent to participate in this study.

### Recruitment

Recruitment is still ongoing in the 6 international study sites. As of March 2025, a total of 195 participants had been recruited, including 48 CTR, 55 PGMC and 92 symptomatic patients (**Fig. 2** and **Table 3**).

**Fig. 2.**
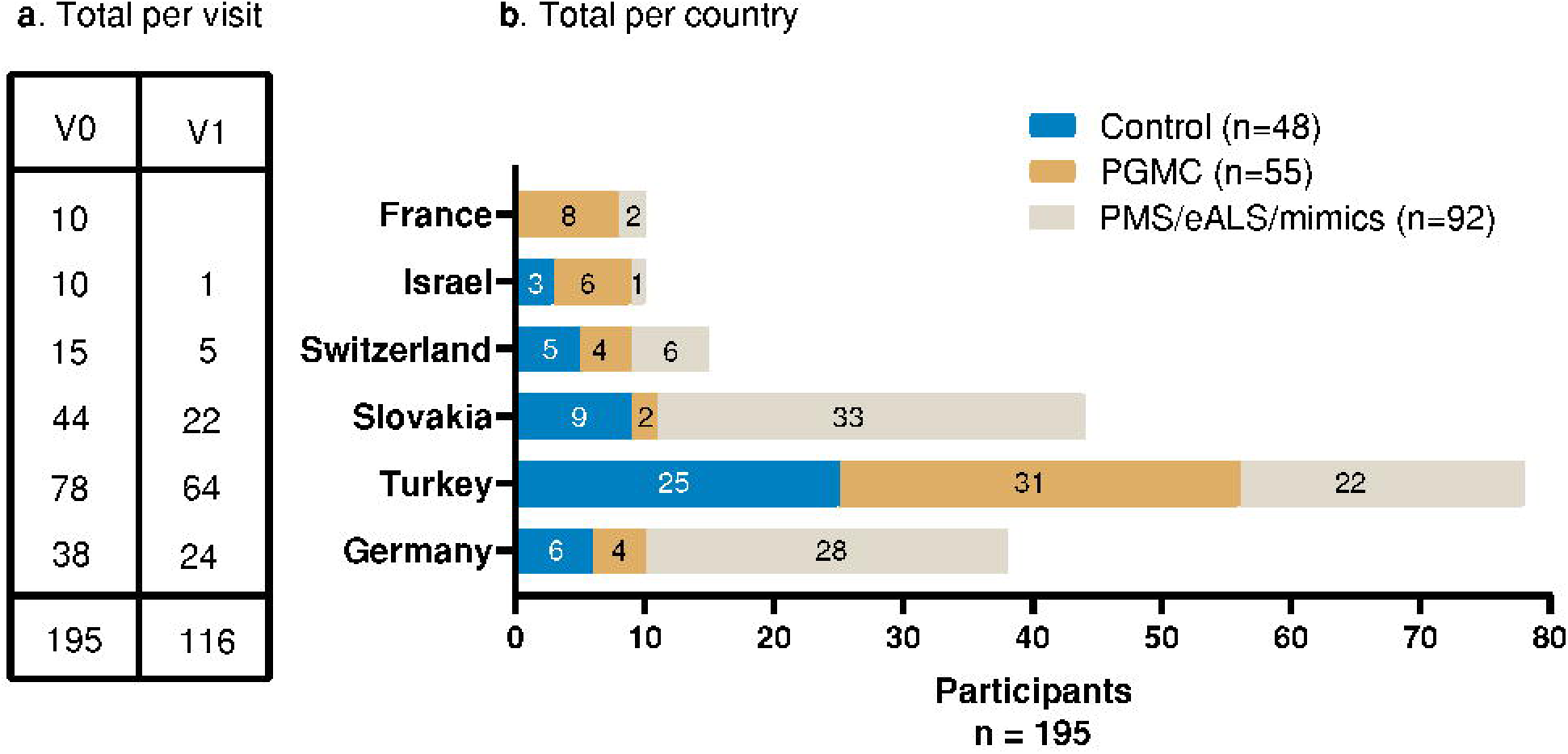
premodiALS recruitment status (as of 03/2025). (a) Number of participants recruited per country in the V0 and V1 for all groups. (b) Number of participants recruited per country in the control, PGMC and the symptomatic patient groups. V: visit; PGMC: pre-symptomatic gene mutation carrier; SYMP: symptomatic patients’ group (early ALS or ALS mimics or pure motor syndrome). *Created in Graphpad Prism. Tzeplaeff, L. (2025)*

**Table 3.**
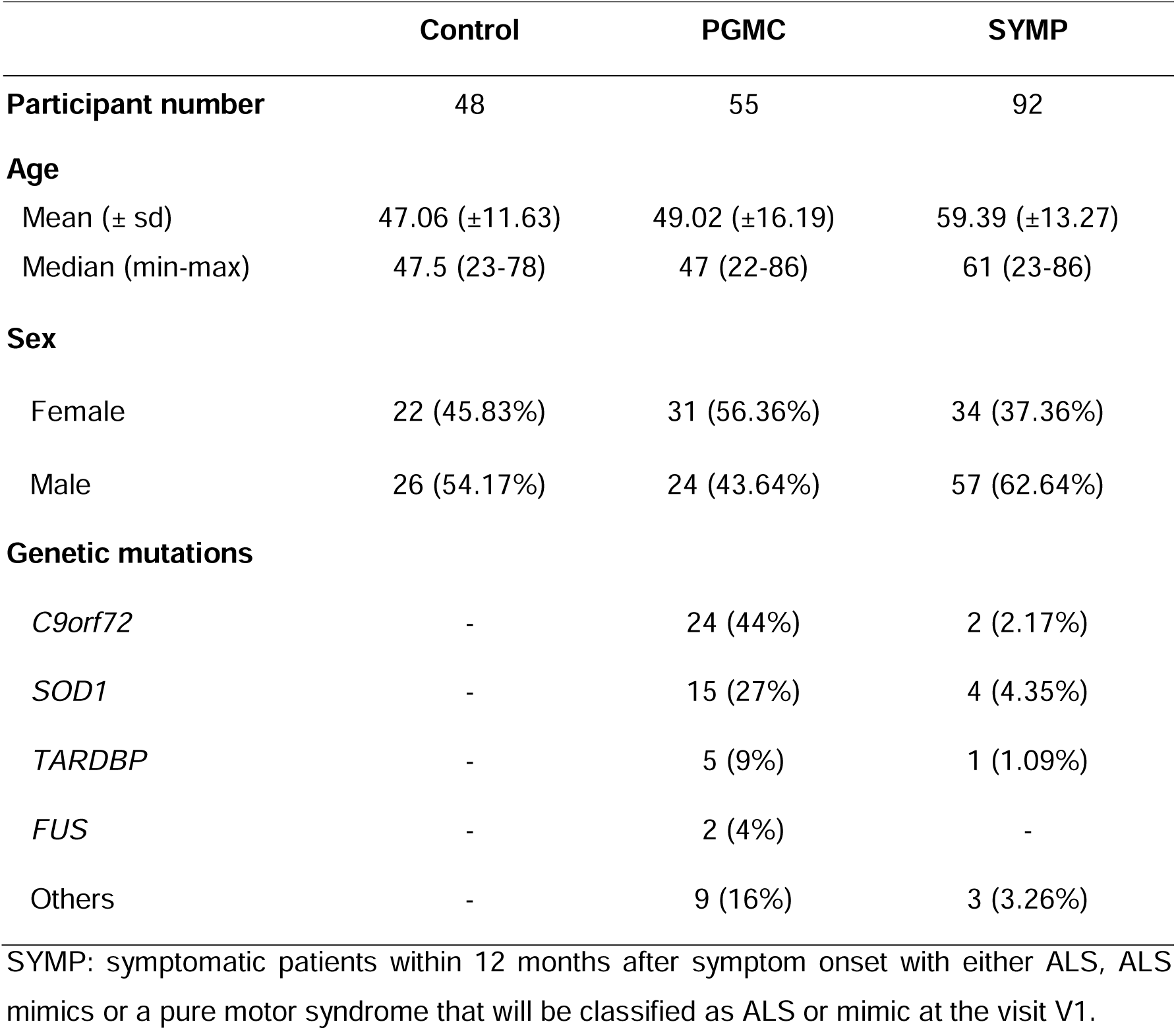
Current cohort description.

To disseminate the design and results of the study, a website has been created, which is updated by regular news sections: https://www.premodials.mh.tum.de/de Scientific results will be disseminated in peer-reviewed, international journals, and at national and international conferences respecting the privacy of the participants.

## Discussion

ALS is mainly diagnosed after irreversible neurological damage has occurred. Although current disease-modifying treatments are still insufficient, earlier and more precise diagnosis could result in earlier intervention, improved disease management, and a better quality of life for patients. Therefore, there is an urgent need for an early diagnosis of the disease. Reducing the diagnosis time from initial symptom presentation to confirmed disease or even predicting the occurrence of phenoconversion in PGMC would be a considerable advance for those affected by ALS and their families. We assume that PGMC gradually and cumulatively develop molecular changes that eventually result in ALS symptoms. Thus, these changes could be visible when comparing the baseline visit (V0) and follow-up visit (V1) of PGMC, and should be unchanged in our CTR cohort. The identification of a presymptomatic signature of ALS would have significant advantages:

1. **Early diagnosis**: The identification of a panel of presymptomatic biomarkers might enable the identification of individuals at risk of phenoconverting to clinically manifest ALS. This could lead to earlier diagnosis and intervention.
2. **Opportunities for early intervention**: The identification of risk biomarkers could facilitate the development of experimental therapies designed to delay or prevent clinically manifest ALS, as exemplified by the ATLAS trial [51]. Additionally, lifestyle modifications could also contribute to delay the apparition of clinically manifest ALS, as discussed in recent guidelines for the clinical management of individuals at elevated genetic risk for ALS [52].
3. **Facilitation of clinical trials**: Identifying individuals at high risk of developing ALS could streamline the recruitment process for clinical trials testing disease-modifying therapies. One example is the ATLAS study, where NfL is used as biomarker evidence of disease activity in presymptomatic SOD1 variant carriers to initiate Tofersen treatment [51]. This could accelerate the development and evaluation of potential treatments for ALS by ensuring that trials enroll participants most likely to benefit.
4. **Personalized medicine**: Biomarkers could enable personalized treatment approaches tailored to individual patients based on their risk profile and disease progression trajectory. This could lead to more effective and targeted therapies, minimizing adverse effects and optimizing treatment outcomes.
5. **Insights into disease pathogenesis**: Understanding the longitudinal behavior of presymptomatic biomarkers could provide valuable insights into the underlying mechanisms and progression of ALS. This knowledge could inform future research efforts aimed at unraveling the complexities of the disease and developing more targeted therapeutic strategies.

With this study we aim to describe a clinico-molecular fingerprint and to provide supervised machine learning models capable of differentiating presymptomatic subjects and patients with ALS from ALS mimics and controls. This could facilitate early diagnosis and treatment and provide further insights into the disease mechanisms of this devastating neurodegenerative condition. One of the limitations of the study is the short follow up period of one year. However, given the successful recruitment so far, we aim to extend the follow-up for another three years, including additional visits V2, V3 and V4 at yearly intervals. We assume that ALS develops through gradual molecular changes and not according to the punctuated equilibrium principle (i.e., motor dysfunction develops suddenly and without antecedent) [52, 53]. However, if PGMC are many years from phenoconversion, these changes may not yet be apparent. In this case we will focus on emerging ALS cases within the PGMC cohort (phenoconverters). Comparing the baseline visit (V0) and follow-up visit (V1) of phenoconverters will identify key features of ALS phenoconversion.

Although premodiALS is a non-interventional, prospective observational study, subjects participating in the study may benefit from data obtained during the visits. As an example, the results of the olfactory testing can be communicated directly to the participants, so that a further evaluation of a possibly existing olfactory disorder can take place if desired. Furthermore, participants will receive a detailed neurological examination, allowing pathological changes to be detected and thus treated earlier, even if originating from other causes. The results of the biomaterials data will not be communicated to the participants, as they are only carried out at a later date and are usually only analyzed and meaningful as a group comparison. However, the study may identify a signature that could be used in the future to prioritize PGMC for a clinical trial of disease-modifying therapies.

Finally, the premodiALS protocol presents a reasonable burden with a low risk of post-interventional bleeding or infection (sampling of blood or CSF), or short-term corneal irritation and foreign body sensation (tear fluid collection) for study participants. Furthermore, the time required for participation (approximately 3 hours per visit) is considered moderate in view of the data and biomaterials collected. While some biomaterial will exclusively be allocated for analyses predetermined in the premodiALS project, others will be biobanked and available for collaborative projects and future analyses. The premodiALS project presented here will provide a copious data source that could be compared to similar initiatives, such as *Pre-fALS*, GENFI or the EPIC studies [20, 26, 54–58]. With the premodiALS protocol, we therefore also aim to promote collaboration, encourage harmonization of protocols across studies to enable comparison of data across larger cohorts, thereby advancing scientific understanding and facilitating more robust research outcomes in the field.

## Data Availability

All data produced in the present work are contained in the manuscript

https://www.premodials.mh.tum.de/de

## Funding

The authors declare financial support was received for the research, authorship, and/or publication of this article. premodiALS is funded by the EU Joint Programme-Neurodegenerative Disease Research (JPND) within the 2021 JPND call for proposals: “Linking pre-diagnosis disturbances of physiological systems to Neurodegenerative Diseases” (premodiALS— https://www.premodials.mh.tum.de/de) PL, LT, MPM and AG are funded by the Bundesministerium für Bildung und Forschung (BMBF: 01ED2204A, 01ED2204B). PL is further supported by the Deutsche Forschungsgemeinschaft (DFG, German Research Foundation) under Germany’s Excellence Strategy within the framework of the Munich Cluster for Systems Neurology (EXC 2145, SyNergy – ID 390857198). MPM is further supported by the European Union’s Horizon 2020 Research and Innovation Programme (Grant agreement No. 950293 - COMBAT-RES). HU received funding from the Scientific and Technological Research Council of Turkey (TÜBİTAK) under the grant number 112N004. MW is funded under the Swiss National Science Foundation (SNSF) (Grant 32ND30_206536). PC and the ALS Center of Tours have been financially supported by the French National Research Agency (ANR) (Grant number: ANR-21-JPW2-0007-03). WK received funding from the Polish National Science Center (NCN) under grant number 2021/03/Y/NZ7/00111. YL was funded by the Ministry of Health of Israel (Grant number: 3-18370). NZ and MTK were funded by the Ministry of Education, Science, Research and Sport of the Slovak Republic. NJA funding was provided by the Swedish Research Council (SRC). MLR (non-funded partner) analytical work is supported by FightMND (05-CIG-Rogers 2022). MB, JW (non-funded partner) and the Pre-fALS study are funded by the U.S. National Institutes of Health (NIH, grant# R01NS105479), the ALS Recovery Fund, and Kimmelman Estate. ANB, VÇ and EB are indebted to Suna and Inan Kıraç Foundation for their generous support of the study and for the inspiring research environment they created.

## Acknowledgments

The authors want to thank all those involved in the design and conduct of this ongoing study. A special thanks goes to Ms. Petra Rau for her support in the preparation of the study. We thank all participants who take part in this study as well as their relatives and caregivers. CL would like to acknowledge the use of resources provided by the University Medical Center Göttingen’s Core Facility Proteomics. ANB, VÇ and EB gratefully acknowledge the use of the services and facilities of KUTTAM

## Statements and Declarations: Competing Interests

The authors declare that they have no conflict of interest.

## Author Contributions

PL developed the study concept, wrote the study protocol. PL and LT coordinate the study, wrote and edited the manuscript. AK, NJA, MB, PC, WK, CL, YL, PMA, MLR, HU, MW, NZ and MPM provided input on study design, and critically edited the manuscript for content. MPM, CM and AG detailed the statistical aspects of the study in the study protocol, and critically reviewed the manuscript. LCG and JW actively contributed to edited the manuscript. PC, YL, HU, MW, NZ, MTK and PL are lead investigators at participating recruiting study centers. LT, NB, HD, AA, HCK, AGC, SD, MK, SBK, EB, VÇ, UB, FK, AFD, AB, RvH, VG, CK and MM participated in patient recruitment and samples processing. NJA, WK, CL, MLR, ANZ, are lead investigators at participating analytic study centers. AM and FK developed the eCRF of the study protocol. All named authors meet the International Committee of Medical Journal Editors (ICMJE) criteria for authorship for this article, take responsibility for the integrity of the work as a whole, and have given their approval for this version to be published.

## Notes

### Competing Interest Statement

The authors have declared no competing interest.

### Funding Statement

This study was funded by the EU Joint Programme Neurodegenerative Disease Research (JPND) within the 2021 JPND call for proposals: Linking pre-diagnosis disturbances of physiological systems to Neurodegenerative Diseases.

### Author Declarations

Ethics committee of the Technical University of Munich gave ethical approval for this work [2022-520_2-S-SB].

