## Supplemental Files 1 for "Identification of a presymptomatic and early disease signature for Amyotrophic Lateral Sclerosis (ALS): protocol of the premodiALS study"

### Questionnaire premodiALS

**This questionnaire should be completed by a study nurse or investigator together with the subject.**

#### Part A. General aspects

**A1.** Please indicate your Handedness. The Handedness, or dominant hand, defines the hand that is spontaneously preferred in the activity and this with greater speed and skill than the non-dominant hand.

- ☐ Right-hander
- ☐ Left-handed
- ☐ Ambidextrous
- ☐ Other (Comment)

**A2.** Please indicate the size of the city where you have lived the longest in the last 10 years.

- ☐ Village (<1000 inhabitants)
- ☐ country town (1000–5000 inhabitants)
- ☐ Small town (5000–20000 inhabitants)
- ☐ Middle town (20000–100000 inhabitants)
- ☐ Large town (>100000 inhabitants)

**A3.** Does one of your relatives (parents, siblings, aunts, uncles, cousins) suffer from a neurological, neurodegenerative or neuromuscular disease?

- ☐ No
- ☐ Yes, and that is (multiple answers possible):
  - ☐ Father
  - ☐ Mother
  - ☐ Sibling
  - ☐ Aunts or uncles
  - ☐ Grandparents
  - ☐ Cousins
  - ☐ Other relatives (please specify): ......

Type of disease

#### Part B. Symptoms

Please indicate the time of the **first** occurrence for each of the symptoms listed below. Use the following scale for each of the questions: M = month, Y = year

| Symptoms | | Never | in the last M | before 2–12 M | before 2–5 Y | before 6–10 Y | more than 10 Y |
|---|---|---|---|---|---|---|---|
| B1. | Decreased strength / dexterity or muscle wasting of the right arm / right hand | ☐ | ☐ | ☐ | ☐ | ☐ | ☐ |
| B2. | Decreased strength / dexterity or muscle wasting left arm / left hand | ☐ | ☐ | ☐ | ☐ | ☐ | ☐ |
| B3. | Decreased strength / dexterity or muscle wasting right leg / foot | ☐ | ☐ | ☐ | ☐ | ☐ | ☐ |
| B4. | Decreased strength / dexterity or muscle wasting of the left leg / foot | ☐ | ☐ | ☐ | ☐ | ☐ | ☐ |
| B5. | Stiffness (spasticity) right arm / right hand | ☐ | ☐ | ☐ | ☐ | ☐ | ☐ |
| B6. | Stiffness (spasticity) left arm / left hand | ☐ | ☐ | ☐ | ☐ | ☐ | ☐ |
| B7. | Stiffness (spasticity) right leg / foot | ☐ | ☐ | ☐ | ☐ | ☐ | ☐ |
| B8. | Stiffness (spasticity) left leg / left foot | ☐ | ☐ | ☐ | ☐ | ☐ | ☐ |
| B9. | Weakness of the neck muscles, problems keeping the head upright | ☐ | ☐ | ☐ | ☐ | ☐ | ☐ |
| B10. | Weakness of the trunk muscles. Standing / walking is only possible with effort | ☐ | ☐ | ☐ | ☐ | ☐ | ☐ |
| B11. | Weakness of the abdominal muscles. Straightening from the lying position to the sitting position is difficult | ☐ | ☐ | ☐ | ☐ | ☐ | ☐ |
| B12. | Trunk bend without support from the arms is only possible with difficulty or not at all | ☐ | ☐ | ☐ | ☐ | ☐ | ☐ |
| B13. | Change of language, e.g. indistinct, muffled, nasal (please also ask relatives) | ☐ | ☐ | ☐ | ☐ | ☐ | ☐ |
| B14. | Difficulty swallowing, e.g. frequent swallowing, swallowing, change in diet, e.g. Mashed | ☐ | ☐ | ☐ | ☐ | ☐ | ☐ |
| B15. | Difficulty breathing / shortness of breath | ☐ | ☐ | ☐ | ☐ | ☐ | ☐ |
| B16. | Muscle tremors / twitches | ☐ | ☐ | ☐ | ☐ | ☐ | ☐ |
| B17. | Increased muscle spasms | ☐ | ☐ | ☐ | ☐ | ☐ | ☐ |
| B18. | Deterioration of memory (please also ask relatives) | ☐ | ☐ | ☐ | ☐ | ☐ | ☐ |
| B19. | Change of personality / behavior, e.g. Irritability, apathy, social withdrawal | ☐ | ☐ | ☐ | ☐ | ☐ | ☐ |
| B20. | Excessive and/or inappropriate emotional outbursts, such as uncontrolled crying/laughing /smiling/yawning | ☐ | ☐ | ☐ | ☐ | ☐ | ☐ |
| B21. | Deterioration of vision | ☐ | ☐ | ☐ | ☐ | ☐ | ☐ |
| B22. | Deterioration of hearing | ☐ | ☐ | ☐ | ☐ | ☐ | ☐ |
| B23. | Deterioration of the sense of smell | ☐ | ☐ | ☐ | ☐ | ☐ | ☐ |
| B24. | Deterioration of the sense of taste | ☐ | ☐ | ☐ | ☐ | ☐ | ☐ |
| B25. | Diarrhea | ☐ | ☐ | ☐ | ☐ | ☐ | ☐ |
| B26. | Fecal incontinence (unable to maintain stool urge) | ☐ | ☐ | ☐ | ☐ | ☐ | ☐ |

| Symptoms | | Never | in the last M | before 2–12 M | before 2–5 Y | before 6–10 Y | more than 10 Y |
|---|---|---|---|---|---|---|---|
| B27. | Constipation (bowel movements less than three times a week) | ☐ | ☐ | ☐ | ☐ | ☐ | ☐ |
| B28. | Feeling of incomplete bowel evacuation after defecation (bowel movements) | ☐ | ☐ | ☐ | ☐ | ☐ | ☐ |
| B29. | Vomiting or nausea | ☐ | ☐ | ☐ | ☐ | ☐ | ☐ |
| B30. | Sudden strong urge to urinate, sometimes with involuntary urine leakage | ☐ | ☐ | ☐ | ☐ | ☐ | ☐ |
| B31. | Feeling of incomplete emptying of the bladder after urination | ☐ | ☐ | ☐ | ☐ | ☐ | ☐ |
| B32. | Urge to urinate with emptying of a small amount of urine | ☐ | ☐ | ☐ | ☐ | ☐ | ☐ |
| B33. | Increased urine excretion | ☐ | ☐ | ☐ | ☐ | ☐ | ☐ |
| B34. | Delayed onset of urine excretion or attenuated urine stream | ☐ | ☐ | ☐ | ☐ | ☐ | ☐ |
| B35. | Urinating more than twice a night | ☐ | ☐ | ☐ | ☐ | ☐ | ☐ |
| B36. | Feeling dizzy or bloodless in the head when getting up from sitting or lying down or after standing for a long time | ☐ | ☐ | ☐ | ☐ | ☐ | ☐ |
| B37. | Low blood pressure | ☐ | ☐ | ☐ | ☐ | ☐ | ☐ |
| B38. | Slow heart rate | ☐ | ☐ | ☐ | ☐ | ☐ | ☐ |
| B39. | Palpitations / tachycardia (rapid heartbeat) | ☐ | ☐ | ☐ | ☐ | ☐ | ☐ |
| B40. | Increased sweating (but not associated with hot weather) | ☐ | ☐ | ☐ | ☐ | ☐ | ☐ |
| B41. | Increased sweating of a body region (e.g. face, arm) | ☐ | ☐ | ☐ | ☐ | ☐ | ☐ |
| B42. | Reduced sweating | ☐ | ☐ | ☐ | ☐ | ☐ | ☐ |
| B43. | Increased freezing (not associated with cold weather) | ☐ | ☐ | ☐ | ☐ | ☐ | ☐ |
| B44. | Excessive sebum secretion of the skin on the face (oily skin) | ☐ | ☐ | ☐ | ☐ | ☐ | ☐ |
| B45. | Decreased sensitivity to touch or temperature in arms and/or legs | ☐ | ☐ | ☐ | ☐ | ☐ | ☐ |
| B46. | Discomfort (e.g. Tingling) in arms and / or legs | ☐ | ☐ | ☐ | ☐ | ☐ | ☐ |
| B47. | Unexplained pain (but not as a result of known diseases / symptoms such as migraines) | ☐ | ☐ | ☐ | ☐ | ☐ | ☐ |
| B48. | Itchy areas of the body | ☐ | ☐ | ☐ | ☐ | ☐ | ☐ |
| B49. | Cold, pale or bluish discolored extremities (arms / legs) | ☐ | ☐ | ☐ | ☐ | ☐ | ☐ |
| B50. | Forgetting things that have been said or happened recently | ☐ | ☐ | ☐ | ☐ | ☐ | ☐ |
| B51. | Forgetting to do everyday things (e.g. taking pills or forgetting to turn off household appliances) | ☐ | ☐ | ☐ | ☐ | ☐ | ☐ |
| B52. | Difficulty concentrating or staying alert for a long period of time (e.g. when reading or speaking, losing thread) | ☐ | ☐ | ☐ | ☐ | ☐ | ☐ |

| Symptoms | | Never | in the last M | before 2–12 M | before 2–5 Y | before 6–10 Y | more than 10 Y |
|---|---|---|---|---|---|---|---|
| B53. | Loss of interest in things that are happening around you (e.g. Conversations) or activities (e.g. Hobbies) | ☐ | ☐ | ☐ | ☐ | ☐ | ☐ |
| B54. | Feeling of sadness, depression or melancholy | ☐ | ☐ | ☐ | ☐ | ☐ | ☐ |
| B55. | Increased irritability, anger or aggressiveness | ☐ | ☐ | ☐ | ☐ | ☐ | ☐ |
| B56. | Nervousness or anxiety in situations that don't usually trigger anxiety | ☐ | ☐ | ☐ | ☐ | ☐ | ☐ |
| B57. | Feelings of panic in situations that usually don't cause panic | ☐ | ☐ | ☐ | ☐ | ☐ | ☐ |
| B58. | Apathy, without the usual "highs" and "lows" | ☐ | ☐ | ☐ | ☐ | ☐ | ☐ |
| B59. | Lack of joy in situations that are usually fun | ☐ | ☐ | ☐ | ☐ | ☐ | ☐ |
| B60. | Seeing or hearing things that you know, or others say, are not there | ☐ | ☐ | ☐ | ☐ | ☐ | ☐ |
| B61. | Believing things happen to you that you know, or others say, are not true | ☐ | ☐ | ☐ | ☐ | ☐ | ☐ |
| B62. | Difficulty making decisions | ☐ | ☐ | ☐ | ☐ | ☐ | ☐ |
| B63. | Irresistible urge or impulse to perform actions that harm you or the general public (e.g. Gambling addiction) | ☐ | ☐ | ☐ | ☐ | ☐ | ☐ |
| B64. | Difficulty falling asleep in the evening | ☐ | ☐ | ☐ | ☐ | ☐ | ☐ |
| B65. | Difficulty sleeping through the night | ☐ | ☐ | ☐ | ☐ | ☐ | ☐ |
| B66. | Vivid / intense dreams or dreams with aggressive content that trigger anxiety | ☐ | ☐ | ☐ | ☐ | ☐ | ☐ |
| B67. | Loud speaking, screaming, scolding or laughing in your sleep | ☐ | ☐ | ☐ | ☐ | ☐ | ☐ |
| B68. | Movements during sleep, as if you were "living out" the content of your dreams | ☐ | ☐ | ☐ | ☐ | ☐ | ☐ |
| B69. | Urge to move the legs and / or unpleasant sensations (e.g. Tingling, pulling), which occur almost exclusively at night or at rest and improve when you move or massage your legs. | ☐ | ☐ | ☐ | ☐ | ☐ | ☐ |
| B70. | Shift of the day-night rhythm: formerly "morning person" now "evening person" | ☐ | ☐ | ☐ | ☐ | ☐ | ☐ |
| B71. | Shift of the day-night rhythm: formerly "evening person" now "morning person” | ☐ | ☐ | ☐ | ☐ | ☐ | ☐ |
| B72. | Difficulty staying awake during activities such as work, driving, or eating. | ☐ | ☐ | ☐ | ☐ | ☐ | ☐ |
| B73. | Extreme form of fatigue (uncontrollable state of fatigue, exhaustion and and lack of energy, regardless of clear external causes) | ☐ | ☐ | ☐ | ☐ | ☐ | ☐ |
| B74. | Morning headaches | ☐ | ☐ | ☐ | ☐ | ☐ | ☐ |
| B75. | Decreased hair growth in the armpits, pubic area and eyes | ☐ | ☐ | ☐ | ☐ | ☐ | ☐ |

| Symptoms | Never | in the last M | before 2–12 M | before 2–5 Y | before 6–10 Y | more than 10 Y |
|---|---|---|---|---|---|---|
| B76. Skin pallor (decreased pigmentation) | ☐ | ☐ | ☐ | ☐ | ☐ | ☐ |
| B77. Increased desire for salt | ☐ | ☐ | ☐ | ☐ | ☐ | ☐ |
| B78. Reduction of the feeling of thirst | ☐ | ☐ | ☐ | ☐ | ☐ | ☐ |
| B79. Increase in the feeling of thirst | ☐ | ☐ | ☐ | ☐ | ☐ | ☐ |
| B80. Sudden sweating / restlessness / tremor | ☐ | ☐ | ☐ | ☐ | ☐ | ☐ |
| B81. Loss of libido (decreased sexual desire) | ☐ | ☐ | ☐ | ☐ | ☐ | ☐ |
| B82. Reduction of appetite | ☐ | ☐ | ☐ | ☐ | ☐ | ☐ |
| B83. Increase in libido (stronger sexual desire) | ☐ | ☐ | ☐ | ☐ | ☐ | ☐ |
| B84. Increase in appetite | ☐ | ☐ | ☐ | ☐ | ☐ | ☐ |
| B85. Frequent falls backwards | ☐ | ☐ | ☐ | ☐ | ☐ | ☐ |
| B86. Problems with balance | ☐ | ☐ | ☐ | ☐ | ☐ | ☐ |
| B87. Erectile dysfunction or impaired arousal | ☐ | ☐ | ☐ | ☐ | ☐ | ☐ |
| B88. Tremor in the arms or legs | ☐ | ☐ | ☐ | ☐ | ☐ | ☐ |
| B89. Frequent yawning (not associated with fatigue, difficult to control at times) | ☐ | ☐ | ☐ | ☐ | ☐ | ☐ |
| B90. Frequent sneezing or sudden sneezing (without a known allergy or cold) | ☐ | ☐ | ☐ | ☐ | ☐ | ☐ |
| B91. Dry eyes | ☐ | ☐ | ☐ | ☐ | ☐ | ☐ |
| B92. Burning or irritated eyes | ☐ | ☐ | ☐ | ☐ | ☐ | ☐ |
| B93. Increased eye tears | ☐ | ☐ | ☐ | ☐ | ☐ | ☐ |
| B94. Excess saliva in the mouth | ☐ | ☐ | ☐ | ☐ | ☐ | ☐ |
| B95. Frequent nasal congestion | ☐ | ☐ | ☐ | ☐ | ☐ | ☐ |
| B96. Frequent "runny" nose | ☐ | ☐ | ☐ | ☐ | ☐ | ☐ |
| B97. Tongue burning | ☐ | ☐ | ☐ | ☐ | ☐ | ☐ |

#### Part C. Contact with the health system

When have you visited doctors / medical facilities (outpatient or inpatient) in the past?

**Multiple crosses for one specialist are possible**, including **the first and the last** visit.

Please do not specify if the visit was only for preventive reasons, i.e. as a precaution.

| Symptoms | | Never | in the last M | before 2–12 M | before 2–5 Y | before 6–10 Y | more than 10 Y |
|---|---|---|---|---|---|---|---|
| C1. | Specialist in Internal Medicine (Gastroenterology) | ☐ | ☐ | ☐ | ☐ | ☐ | ☐ |
| C2. | Specialist in Internal Medicine (Cardiology) | ☐ | ☐ | ☐ | ☐ | ☐ | ☐ |
| C3. | Specialist in Internal Medicine (Pulmonology) | ☐ | ☐ | ☐ | ☐ | ☐ | ☐ |
| C4. | Specialist in Internal Medicine (Endocrinology) | ☐ | ☐ | ☐ | ☐ | ☐ | ☐ |
| C5. | Specialist in Internal Medicine (Hematology / Oncology) | ☐ | ☐ | ☐ | ☐ | ☐ | ☐ |
| C6. | Specialist in Internal Medicine (Nephrology) | ☐ | ☐ | ☐ | ☐ | ☐ | ☐ |
| C7. | Specialist in Internal Medicine (Rheumatology) | ☐ | ☐ | ☐ | ☐ | ☐ | ☐ |
| C8. | Specialist in Neurology | ☐ | ☐ | ☐ | ☐ | ☐ | ☐ |
| C9. | Specialist in Psychiatry | ☐ | ☐ | ☐ | ☐ | ☐ | ☐ |
| C10. | Specialist in ENT (ear, nose and throat) | ☐ | ☐ | ☐ | ☐ | ☐ | ☐ |
| C11. | Specialist in Ophthalmology | ☐ | ☐ | ☐ | ☐ | ☐ | ☐ |
| C12. | Specialist in Gynecology | ☐ | ☐ | ☐ | ☐ | ☐ | ☐ |
| C13. | Specialist in Urology | ☐ | ☐ | ☐ | ☐ | ☐ | ☐ |
| C14. | Specialist in Surgery / Orthopedics | ☐ | ☐ | ☐ | ☐ | ☐ | ☐ |
| C15. | Specialist in Dermatology | ☐ | ☐ | ☐ | ☐ | ☐ | ☐ |
| C16. | Specialist in Psychosomatics | ☐ | ☐ | ☐ | ☐ | ☐ | ☐ |
| C17. | Physiotherapy | ☐ | ☐ | ☐ | ☐ | ☐ | ☐ |
| C18. | Occupational therapy | ☐ | ☐ | ☐ | ☐ | ☐ | ☐ |
| C19. | Speech therapy | ☐ | ☐ | ☐ | ☐ | ☐ | ☐ |
| C20. | Psychotherapy | ☐ | ☐ | ☐ | ☐ | ☐ | ☐ |
| C21. | Healer | ☐ | ☐ | ☐ | ☐ | ☐ | ☐ |
| C22. | Other, please specify | ☐ | ☐ | ☐ | ☐ | ☐ | ☐ |

Other:

#### Part D. Pre-existing conditions

Are you currently suffering or have you suffered from diseases from subsequent areas in the past?

Please indicate when the **first** symptoms appeared or, if the condition did not cause any symptoms, when it was diagnosed.

When answering the question, please also think about your medication, e.g. medication for high blood pressure → high blood pressure is a disease of the cardiovascular system.

| Pre-existing conditions | Never | in the last M | before 2–12 M | before 2–5 Y | before 6–10 Y | more than 10 Y |
|---|---|---|---|---|---|---|
| D1. **Which disease of the cardiovascular system is or was present and since when?** | | | | | | |
| 1. Hypertension | ☐ | ☐ | ☐ | ☐ | ☐ | ☐ |
| 2. Heart attack | ☐ | ☐ | ☐ | ☐ | ☐ | ☐ |
| 3. Coronary heart disease (narrowing of the coronary arteries) | ☐ | ☐ | ☐ | ☐ | ☐ | ☐ |
| 4. Peripheral arterial occlusive disease of the extremities (PAD) | ☐ | ☐ | ☐ | ☐ | ☐ | ☐ |
| 5. Other, please specify..... | ☐ | ☐ | ☐ | ☐ | ☐ | ☐ |
| D2. **Which disease of the respiratory system is or was present and when?** | | | | | | |
| 1. Pneumonia | ☐ | ☐ | ☐ | ☐ | ☐ | ☐ |
| 2. Hay fever | ☐ | ☐ | ☐ | ☐ | ☐ | ☐ |
| 3. Chronic sinusitis | ☐ | ☐ | ☐ | ☐ | ☐ | ☐ |
| 4. Chronic obstructive pulmonary disease (COPD) | ☐ | ☐ | ☐ | ☐ | ☐ | ☐ |
| 5. Bronchial asthma | ☐ | ☐ | ☐ | ☐ | ☐ | ☐ |
| 6. Other, please specify..... | ☐ | ☐ | ☐ | ☐ | ☐ | ☐ |
| D3. **Which disease of the digestive system is or was present and when?** | | | | | | |
| 1. Reflux disease | ☐ | ☐ | ☐ | ☐ | ☐ | ☐ |
| 2. Stomach or duodenal ulcer | ☐ | ☐ | ☐ | ☐ | ☐ | ☐ |
| 3. Chronic inflammation of the gastric mucosa | ☐ | ☐ | ☐ | ☐ | ☐ | ☐ |
| 4. Appendicitis | ☐ | ☐ | ☐ | ☐ | ☐ | ☐ |
| 5. Hernia | ☐ | ☐ | ☐ | ☐ | ☐ | ☐ |
| 6. Crohn's disease | ☐ | ☐ | ☐ | ☐ | ☐ | ☐ |
| 7. Ulcerative colitis | ☐ | ☐ | ☐ | ☐ | ☐ | ☐ |
| 8. Coeliac disease | ☐ | ☐ | ☐ | ☐ | ☐ | ☐ |
| 9. Intestinal obstruction (ileus) | ☐ | ☐ | ☐ | ☐ | ☐ | ☐ |
| 10. Diverticulosis | ☐ | ☐ | ☐ | ☐ | ☐ | ☐ |
| 11. Irritable bowel syndrome | ☐ | ☐ | ☐ | ☐ | ☐ | ☐ |
| 12. Hamorrhoids | ☐ | ☐ | ☐ | ☐ | ☐ | ☐ |
| 13. Pancreatitis | ☐ | ☐ | ☐ | ☐ | ☐ | ☐ |
| 14. Cirrhosis | ☐ | ☐ | ☐ | ☐ | ☐ | ☐ |
| 15. Cholelithiasis (gallstones) | ☐ | ☐ | ☐ | ☐ | ☐ | ☐ |
| 16. Other, please specify..... | ☐ | ☐ | ☐ | ☐ | ☐ | ☐ |

| Pre-existing conditions | Never | in the last M | before 2–12 M | before 2–5 Y | before 6–10 Y | more than 10 Y |
|---|---|---|---|---|---|---|
| D4. **Which disease of the nervous system is or was present and since when?** | | | | | | |
| 1. Stroke | ☐ | ☐ | ☐ | ☐ | ☐ | ☐ |
| 2. Headache | ☐ | ☐ | ☐ | ☐ | ☐ | ☐ |
| 3. Parkinson's syndrome | ☐ | ☐ | ☐ | ☐ | ☐ | ☐ |
| 4. Multiple sclerosis | ☐ | ☐ | ☐ | ☐ | ☐ | ☐ |
| 5. Epilepsy | ☐ | ☐ | ☐ | ☐ | ☐ | ☐ |
| 6. Polyneuropathy | ☐ | ☐ | ☐ | ☐ | ☐ | ☐ |
| 7. Meningitis | ☐ | ☐ | ☐ | ☐ | ☐ | ☐ |
| 8. Other, please specify..... | ☐ | ☐ | ☐ | ☐ | ☐ | ☐ |
| D5. **Which disease of the psyche or behavioral disorder is or was present and when?** | | | | | | |
| 1. Dementia (e.g. Alzheimer's dementia) | ☐ | ☐ | ☐ | ☐ | ☐ | ☐ |
| 2. Alcoholism | ☐ | ☐ | ☐ | ☐ | ☐ | ☐ |
| 3. Depressive disorder | ☐ | ☐ | ☐ | ☐ | ☐ | ☐ |
| 4. Anxiety disorder | ☐ | ☐ | ☐ | ☐ | ☐ | ☐ |
| 5. Panic disorder | ☐ | ☐ | ☐ | ☐ | ☐ | ☐ |
| 6. Somatoform diseases (psychosomatic disease) | ☐ | ☐ | ☐ | ☐ | ☐ | ☐ |
| 7. Schizophrenia | ☐ | ☐ | ☐ | ☐ | ☐ | ☐ |
| 8. Personality disorder | ☐ | ☐ | ☐ | ☐ | ☐ | ☐ |
| 9. Other, please specify..... | ☐ | ☐ | ☐ | ☐ | ☐ | ☐ |
| D6. **Which disease of the hormonal system or the diet or metabolism is or was present and since when?** | | | | | | |
| 1. Hypothyroidism | ☐ | ☐ | ☐ | ☐ | ☐ | ☐ |
| 2. Hyperthyroidism | ☐ | ☐ | ☐ | ☐ | ☐ | ☐ |
| 3. Diabetes mellitus, type 1 | ☐ | ☐ | ☐ | ☐ | ☐ | ☐ |
| 4. Diabetes mellitus, type 2 | ☐ | ☐ | ☐ | ☐ | ☐ | ☐ |
| 5. Hypercholesterolemia or hypertriglyceridemia | ☐ | ☐ | ☐ | ☐ | ☐ | ☐ |
| 6. Lactose intolerance | ☐ | ☐ | ☐ | ☐ | ☐ | ☐ |
| 7. Other, please specify..... | ☐ | ☐ | ☐ | ☐ | ☐ | ☐ |
| D7. **Which disease of the urinary and genital organs is or was present and when?** | | | | | | |
| 1. Chronic renal insufficiency | ☐ | ☐ | ☐ | ☐ | ☐ | ☐ |
| 2. Kidney or ureter stones | ☐ | ☐ | ☐ | ☐ | ☐ | ☐ |
| 3. Infertility | ☐ | ☐ | ☐ | ☐ | ☐ | ☐ |
| 4. Missing, too weak or too rare menstruation | ☐ | ☐ | ☐ | ☐ | ☐ | ☐ |
| 5. Too heavy, too frequent or irregular menstruation | ☐ | ☐ | ☐ | ☐ | ☐ | ☐ |
| 6. Stress incontinence (stress incontinence) | ☐ | ☐ | ☐ | ☐ | ☐ | ☐ |

| Pre-existing conditions | Never | in the last M | before 2–12 M | before 2–5 Y | before 6–10 Y | more than 10 Y |
|---|---|---|---|---|---|---|
| 7. Benign prostatic hyperplasia (benign enlargement of the prostate) | ☐ | ☐ | ☐ | ☐ | ☐ | ☐ |
| 8. Other, please specify..... | ☐ | ☐ | ☐ | ☐ | ☐ | ☐ |
| D8. **Which cancer (malignant neoplasm) is or was present and when?** | | | | | | |
| 1. Prostate cancer | ☐ | ☐ | ☐ | ☐ | ☐ | ☐ |
| 2. Colorectal cancer (colon) | ☐ | ☐ | ☐ | ☐ | ☐ | ☐ |
| 3. Stomach cancer | ☐ | ☐ | ☐ | ☐ | ☐ | ☐ |
| 4. Esophageal cancer | ☐ | ☐ | ☐ | ☐ | ☐ | ☐ |
| 5. Liver cancer | ☐ | ☐ | ☐ | ☐ | ☐ | ☐ |
| 6. Pancreatic cancer | ☐ | ☐ | ☐ | ☐ | ☐ | ☐ |
| 7. Throat cancer | ☐ | ☐ | ☐ | ☐ | ☐ | ☐ |
| 8. Lung cancer | ☐ | ☐ | ☐ | ☐ | ☐ | ☐ |
| 9. Black skin cancer (melanoma) | ☐ | ☐ | ☐ | ☐ | ☐ | ☐ |
| 10. White skin cancer (basal cell carcinoma or squamous cell carcinoma) | ☐ | ☐ | ☐ | ☐ | ☐ | ☐ |
| 11. Breast cancer | ☐ | ☐ | ☐ | ☐ | ☐ | ☐ |
| 12. Cervical cancer | ☐ | ☐ | ☐ | ☐ | ☐ | ☐ |
| 13. Kidney | ☐ | ☐ | ☐ | ☐ | ☐ | ☐ |
| 14. Thyroid cancer | ☐ | ☐ | ☐ | ☐ | ☐ | ☐ |
| 15. Brain tumor | ☐ | ☐ | ☐ | ☐ | ☐ | ☐ |
| 16. Lymphoma | ☐ | ☐ | ☐ | ☐ | ☐ | ☐ |
| 17. Leukemia | ☐ | ☐ | ☐ | ☐ | ☐ | ☐ |
| 18. Other, please specify..... | ☐ | ☐ | ☐ | ☐ | ☐ | ☐ |
| D9. **Which disease of the musculoskeletal system is or was present and when?** | | | | | | |
| 1. Osteoporosis | ☐ | ☐ | ☐ | ☐ | ☐ | ☐ |
| 2. Scoliosis | ☐ | ☐ | ☐ | ☐ | ☐ | ☐ |
| 3. Spinal disc herniation | ☐ | ☐ | ☐ | ☐ | ☐ | ☐ |
| 4. Spinal canal stenosis | ☐ | ☐ | ☐ | ☐ | ☐ | ☐ |
| 5. Bone fractures/fractures | ☐ | ☐ | ☐ | ☐ | ☐ | ☐ |
| 6. Muscle disease/myopathy | ☐ | ☐ | ☐ | ☐ | ☐ | ☐ |
| 7. Other, please specify..... | ☐ | ☐ | ☐ | ☐ | ☐ | ☐ |
| D10. **Which infectious disease is or was present and when?** | | | | | | |
| 1. Tuberculosis | ☐ | ☐ | ☐ | ☐ | ☐ | ☐ |
| 2. Syphilis | ☐ | ☐ | ☐ | ☐ | ☐ | ☐ |
| 3. Gonococcal infection | ☐ | ☐ | ☐ | ☐ | ☐ | ☐ |
| 4. Chlamydia infection | ☐ | ☐ | ☐ | ☐ | ☐ | ☐ |
| 5. Labial Herpes | ☐ | ☐ | ☐ | ☐ | ☐ | ☐ |
| 6. Shingles | ☐ | ☐ | ☐ | ☐ | ☐ | ☐ |
| 7. HIV | ☐ | ☐ | ☐ | ☐ | ☐ | ☐ |

| | Pre-existing conditions | Never | in the last M | before 2–12 M | before 2–5 Y | before 6–10 Y | more than 10 Y |
|---|---|---|---|---|---|---|---|
| 8. | COVID-19 | ☐ | ☐ | ☐ | ☐ | ☐ | ☐ |
| 9. | Other, please specify..... | ☐ | ☐ | ☐ | ☐ | ☐ | ☐ |
| D11. | **Which disease of the skin is or was present and when?** | | | | | | |
| 1. | Neurodermatitis (atopic eczema) | ☐ | ☐ | ☐ | ☐ | ☐ | ☐ |
| 2. | Psoriasis | ☐ | ☐ | ☐ | ☐ | ☐ | ☐ |
| 3. | Other, please specify..... | ☐ | ☐ | ☐ | ☐ | ☐ | ☐ |
| D12. | **Which disease of the ear is or was present and when?** | | | | | | |
| 1. | Hearing impairment | ☐ | ☐ | ☐ | ☐ | ☐ | ☐ |
| 2. | Otitis media | ☐ | ☐ | ☐ | ☐ | ☐ | ☐ |
| 3. | Benign paroxysmal positional vertigo (BPPV) | ☐ | ☐ | ☐ | ☐ | ☐ | ☐ |
| 4. | Other, please specify..... | ☐ | ☐ | ☐ | ☐ | ☐ | ☐ |
| D13. | **Which disease of the eye is or was present and when?** | | | | | | |
| 1. | Myopia | ☐ | ☐ | ☐ | ☐ | ☐ | ☐ |
| 2. | Farsightedness | ☐ | ☐ | ☐ | ☐ | ☐ | ☐ |
| 3. | Presbyopia | ☐ | ☐ | ☐ | ☐ | ☐ | ☐ |
| 4. | Cataract | ☐ | ☐ | ☐ | ☐ | ☐ | ☐ |
| 5. | Glaucoma | ☐ | ☐ | ☐ | ☐ | ☐ | ☐ |
| 6. | Retinal detachment | ☐ | ☐ | ☐ | ☐ | ☐ | ☐ |
| 7. | Other, please specify..... | ☐ | ☐ | ☐ | ☐ | ☐ | ☐ |
| D14. | **Did you have any illnesses during or before pregnancy, childbirth or the puerperium and if so, when?** | | | | | | |
| 1. | Miscarriage | ☐ | ☐ | ☐ | ☐ | ☐ | ☐ |
| 2. | Insufficient milk production during lactation | ☐ | ☐ | ☐ | ☐ | ☐ | ☐ |
| 3. | Other, please specify..... | ☐ | ☐ | ☐ | ☐ | ☐ | ☐ |
| D15. | **Which rheumatic or autoimmune disease is or was present and when?** | | | | | | |
| 1. | Rheumatoid arthritis | ☐ | ☐ | ☐ | ☐ | ☐ | ☐ |
| 2. | Lupus erythematosus | ☐ | ☐ | ☐ | ☐ | ☐ | ☐ |
| 3. | Raynaud's syndrome | ☐ | ☐ | ☐ | ☐ | ☐ | ☐ |
| 4. | Scleroderma | ☐ | ☐ | ☐ | ☐ | ☐ | ☐ |
| 5. | Sjögren's syndrome | ☐ | ☐ | ☐ | ☐ | ☐ | ☐ |
| 6. | Other, please specify..... | ☐ | ☐ | ☐ | ☐ | ☐ | ☐ |
| D16. | **Have you ever received a blood transfusion?** | | | | | | |
| 1. | Receiving blood transfusion | ☐ | ☐ | ☐ | ☐ | ☐ | ☐ |

#### Part E. Nutrition and weight

| | |
|---|---|
| E1. Please indicate your **height** | cm |
| E2. Please enter your **current weight** | kg |
| E3. Please indicate your **weight 1 year ago** | kg |
| E4. Please indicate your **weight 5 years ago** | kg |
| E5. Please indicate your **weight 10 years ago** | kg |

| Nutrition / Diets | Never | in the last M | before 2–12 M | before 2–5 Y | before 6–10 Y | more than 10 Y |
|---|---|---|---|---|---|---|
| E6. Have you followed diets in the form of reduced calorie intake (for weight loss)? | ☐ | ☐ | ☐ | ☐ | ☐ | ☐ |
| E7. Have you ever fasted in the past? | ☐ | ☐ | ☐ | ☐ | ☐ | ☐ |
| E8. Have you changed your diet to a vegetarian diet in the past? | ☐ | ☐ | ☐ | ☐ | ☐ | ☐ |
| E9. Have you changed your diet to a vegan diet in the past? | ☐ | ☐ | ☐ | ☐ | ☐ | ☐ |
| E10. Have you changed your diet to a lactose-free diet in the past? | ☐ | ☐ | ☐ | ☐ | ☐ | ☐ |
| E11. Have you changed your diet to a fructose-free diet in the past? | ☐ | ☐ | ☐ | ☐ | ☐ | ☐ |
| E12. Have you changed your diet to a gluten-free diet in the past? | ☐ | ☐ | ☐ | ☐ | ☐ | ☐ |
| E13. Do you regularly take dietary supplements (e.g. protein, vitamins or others)? | ☐ | ☐ | ☐ | ☐ | ☐ | ☐ |
| E14. Other dietary changes, please explain: ...... | ☐ | ☐ | ☐ | ☐ | ☐ | ☐ |

#### Part F. Lifestyle, social aspects, special events

F1. What is your current partnership status?
- ☐ Single
- ☐ Living in partnership
- ☐ Other (Comment)

F2. Do you have children ?
- ☐ Yes
- ☐ No

F3. How many children do you have?

| |
|---|
| Number of children |

F4. How many years of school education did you have?

| |
|---|
| Years of school education |

F5. What best describes your highest professional degree?
- ☐ No professional qualification
- ☐ Apprenticeship / Skilled worker qualification
- ☐ Technical college / technical school / commercial academy
- ☐ Bachelor's Degree
- ☐ Master's Degree
- ☐ Doctorate

Have there been any subsequent changes in your private or professional life in the past?

| Changes professionally or privately | Never | in the last M | before 2–12 M | before 2–5 Y | before 6–10 Y | more than 10 Y |
|---|---|---|---|---|---|---|
| F6. Change of profession | ☐ | ☐ | ☐ | ☐ | ☐ | ☐ |
| F7. Change of employer | ☐ | ☐ | ☐ | ☐ | ☐ | ☐ |
| F8. Termination of employment | ☐ | ☐ | ☐ | ☐ | ☐ | ☐ |
| F9. Work-related relocation | ☐ | ☐ | ☐ | ☐ | ☐ | ☐ |
| F10. Death of a very close person, e.g. partner, relative | ☐ | ☐ | ☐ | ☐ | ☐ | ☐ |
| F11. Serious illness of a very close person, e.g. partner, relative | ☐ | ☐ | ☐ | ☐ | ☐ | ☐ |
| F12. Termination of a partnership (separation, divorce) | ☐ | ☐ | ☐ | ☐ | ☐ | ☐ |
| F13. Another change in life that you think could have significantly shaped your life or personality. Please specify: ......... | ☐ | ☐ | ☐ | ☐ | ☐ | ☐ |

F14. Do you currently consume caffeinated beverages (at least 1 cup per/d) or have you ever consumed caffeinated beverages regularly in your life?
- ☐ Yes, current
- ☐ Yes, in the past
- ☐ No, never

F15. Please indicate  the average amount of caffeine consumption. (Please estimate approximate quantity.  A cup of black coffee is about the same  as the caffeine content of one liter of cola or a can of energy Drink.)

- ☐ nothing
- ☐ Little (e.g. 1 – 2 cups of coffee per day)
- ☐ Moderate (e.g. 3 – 4 cups of coffee per day)
- ☐ Lots (5 and more cups of coffee per day)

F16. Do you regularly consume alcoholic beverages or have you ever consumed alcoholic beverages regularly in your life ?

- ☐ Yes, current
- ☐ Yes, in the past
- ☐ No, never

F17. How much alcoholic beverages do you consume on average or have you consumed on average?  One standard drink corresponds to: 0.25 l beer or 0.1 l wine/sparkling wine or 0.04 l schnapps

- ☐ Only occasionally
- ☐ Up to 2 standard drinks per day (men) or 1 standard drink per day (women), at max. 5 days a week
- ☐ More than 2 standard drinks per day (men) or 1 standard drink per day (women), at max. 5 days a week

F18. Do you currently smoke or have you ever smoked cigarettes (or cigars etc.) regularly in your life? (Regular = at least 5 cigarettes/cigarillos per week or 2 cigars/pipes per week)

- ☐ Yes, current
- ☐ Yes, in the past
- ☐ No, never

F19. Do you currently use illegal drugs or have you ever used illegal drugs in your life?

- ☐ No, never
- ☐ Yes, currently. Please specify in F20.
- ☐ Yes, in the past. Please specify in F20.

F20. What illegal drugs do you use or have you used?  Multiple answers possible.

- ☐ Marijuana
- ☐ MDMA / Ecstasy
- ☐ Amphetamines (speed, crystal meth etc.)
- ☐ Cocaine
- ☐ LSD
- ☐ Opiates
- ☐ Heroin
- ☐ Fungi
- ☐ Other, please specify:

F21. Have you used psychotropic drugs (e.g., benzodiazepines)?

- ☐ No, never
- ☐ Yes, currently, please specify in F22
- ☐ Yes, in the past, please specify in F22

F22. If yes what psychotropic drugs did you take?

- ☐ benzodiazepines
- ☐ antidepressants
- ☐ Mood stabilizers (phase prophylactics)
- ☐ Antipsychotics (neuroleptics)
- ☐ antidementives
- ☐ Psychostimulants
- ☐ sleeping pills
- ☐ Other, please specify:

F23. Please describe the degree of physical activity in the context of your professional activity. This information relates to the professional activity you have been doing for most of your life.

- ☐ Low (e.g. office job or other sedentary activity)
- ☐ Moderate (e.g. nursing)
- ☐ Intensive (e.g. construction work)

F24. Have you ever exercised regularly in your life?

- ☐ No.
- ☐ Yes, then please specify in F25 and F26.

F25. Describe the level of the physical activity:

- ☐ Low (1-2 hours per week)
- ☐ Moderate (3-4 hours per week)
- ☐ Intensive (5 or more hours per week)

F26. Which sport(s) did you do superficially:

- ☐ Contact sports (e.g. football, boxing, rugby, karate, football, hockey)
- ☐ Limited contact sports (e.g. basketball, handball, volleyball)
- ☐ Endurance sports (e.g. jogging, swimming, cycling)
- ☐ Racket sports (e.g. tennis, badminton, table tennis)
- ☐ Strength sports (e.g. weightlifting, high intensity training)

| Special Events | Never | in the last M | before 2–12 M | before 2–5 Y | before 6–10 Y | more than 10 Y |
|---|---|---|---|---|---|---|
| F27. Have you experienced episodes of exceptional physical activity in the past (e.g. marathon, triathlon, Ironman, competitive sports) | ☐ | ☐ | ☐ | ☐ | ☐ | ☐ |
| F28. Have you ever experienced an electric shock with mains voltage or higher? | ☐ | ☐ | ☐ | ☐ | ☐ | ☐ |
| F29. Have you ever donated blood? | ☐ | ☐ | ☐ | ☐ | ☐ | ☐ |

#### Part G. Special assessment of motor symptoms

The following questions relate to various abilities or symptoms that we are particularly interested in in this study. That doesn't mean you have to have any of these symptoms. If an area is not affected, specify "undisturbed". If a change is only changed by a short-term illness, please indicate your normal state, e.g. if you usually have no problems with speaking, but are hoarse due to a cold, please indicate "undisturbed" at F1.

Give the answer that you think is most appropriate today.  No multiple answers possible.

G1. Speech.

- ☐ Normal speech processes
- ☐ Detectable speech disturbance - Some detectable alteration in speech.
- ☐ Intelligible with repeating - Some repetition is required to understand the speech.
- ☐ Speech combined with nonvocal communication - Gestures or communication aids are required to understand the speech.
- ☐ Loss of useful speech - impossible for the person to communicate verbally.

G2. Salivation

- ☐ Normal.
- ☐ Slight but definite excess of saliva in mouth - may have nighttime drooling.
- ☐ Moderately excessive saliva - may have minimal drooling.
- ☐ Marked excess of saliva with some drooling.
- ☐ Marked drooling - Requires constant tissue or handkerchief.

G3. Swallowing

- ☐ Normal eating habits - No difficulty swallowing, can eat any foods/liquids of choice.
- ☐ Early eating problems, occasional choking - Ask whether patient is careful with any foods because they get caught in his/her throat; can still eat all foods of choice but with occasional choking.
- ☐ Dietary consistency changes - Avoids certain foods or requires that consistency of foods be changed.
- ☐ Needs supplemental tube feeding.
- ☐ Nothing per oral NPO - Exclusively parenteral or enteral feeding.

G4. Handwriting

- ☐ Normal.
- ☐ Slow or sloppy: all words are legible.
- ☐ Not all words are legible.
- ☐ No words are legible, but can still grip pen.
- ☐ Unable to grip pen.

G5. Cut food and handle utensils OR handle feeding tube and accessories

- ☐ Normal.
- ☐ Somewhat slow and clumsy, but no help needed - Some difficulty cutting or handling utensils by methods used prior to disease onset but patient continues to do so independently.
  OR
  Clumsy, but able to perform all manipulations independently.
- ☐ Can cut most foods (>50%), although slow and clumsy; some help needed - Some difficulty cutting or handling utensils by methods used prior to disease onset; patient requires assistance, but still tries to cut some foods, and still does >50% of the task successfully.
  OR
  Some help needed with closures and fasteners.
- ☐ Food must be cut by someone, but can still feed slowly. Patient cannot cut foods by methods used prior to disease onset, but still tries to feed him/herself and succeeds at least occasionally.
  OR
  Provides minimal assistance to caregiver.
- ☐ Needs to be fed OR Unable to perform any aspect of task.

G6. Dressing and hygiene

- ☐ Normal function - Patient has NO difficulty, and is still completely independent in dressing and hygiene.
- ☐ Independent; can complete self-care with effort or decreased efficiency - Patient still completely independent in dressing but requires more effort to dress; no substitute methods are used to dress.
- ☐ Intermittent assistance or substitute methods - Patient requires occasional assistance or the use of assistive devices or substitute methods (e.g., pull-on clothes, Velcro closure or shoes, pre-buttoned shirt, lying down to don pants) in dressing and hygiene. Methods used are now different than those used prior to disease onset.
- ☐ Needs attendant for self-care - Means patient needs daily caregiver assistance with dressing but patient has some level of function.
- ☐ Total dependence.

G7. Turning in bed and adjusting bed clothes

- ☐ Normal function.
- ☐ Somewhat slow and clumsy, but no help needed.
- ☐ Can turn alone, or adjust sheets, but with great difficulty - Patient can turn alone or adjust sheets, but completes task with great difficulty; no help needed.
- ☐ Can initiate, but not turn or adjust sheets alone.
- ☐ Helpless.

G8. Walking

- ☐ Normal.
- ☐ Early ambulation difficulties - Notes some difficulty, but walks without assistance.
- ☐ Walks with assistance - Includes AFO, cane, walker, or a caregiver.
- ☐ Non-ambulatory function movement only -Patient is able to move lower extremities partially for functional movement; able to stand for transfers, but unable to walk.
- ☐ No purposeful leg movement.

G9. Climbing stairs

- ☐ Normal.
- ☐ Slow.
- ☐ Mild unsteadiness or fatigue. Patient needs to rest between steps, or feels unsteady, but does not need rail.
- ☐ Needs assistance. Patient needs assistance including handrail or caregiver.
- ☐ Cannot do.

G10.Dyspnea

- ☐ None.
- ☐ Occurs when walking.
- ☐ Occurs with one or more of the following: eating, bathing, dressing.
- ☐ Occurs at rest: difficulty breathing when either sitting or lying.
- ☐ Significant difficulty: considering using mechanical respiratory support.

G11.Orthopnea

- ☐ None.
- ☐ Some difficulty sleeping at night due to shortness of breath, does not routinely use more than two pillows.
- ☐ Needs extra pillows in order to sleep (more than two).
- ☐ Can only sleep sitting up.

- ☐ Unable to sleep without mechanical assistance.

G12. Respiratory insufficiency

- ☐ None.
- ☐ Intermittent use of BiPAP.
- ☐ Continuous use of BiPAP during the night.
- ☐ Continuous use of BiPAP during day and night.
- ☐ Invasive mechanical ventilation by intubation or tracheostomy.

Thank you for participating in this survey!
